# A Smaller Right Ventricle Results in Poorer Exercise Performance in Adolescents After Surgical Repair of Tetralogy of Fallot

**DOI:** 10.1101/2024.04.12.24305748

**Authors:** Christiane Mhanna, Katerina Kourpas, Takeshi Tsuda

## Abstract

**Background:** Chronic pulmonary valve insufficiency frequently results in right ventricular (RV) dilatation and dysfunction in surgically repaired tetralogy of Fallot (rTOF). Correlations between peak exercise performance and progression of RV remodeling in rTOF remain elusive.

**Methods:** Patients with rTOF were reviewed with cardiopulmonary exercise testing (CPET) and cardiac magnetic resonance (CMR). Peak and submaximal CPET parameters were obtained. Both RV and left ventricular (LV) volume were measured in end-systole (RVESV and LVESV, respectively) and end-diastole (LVEDV and RVEDV, respectively). Stroke volume (SV), ejection fraction (EF), and pulmonary regurgitant fraction (RF) were calculated.

**Results:** Thirty-seven patients (17 ± 5 years; 22 females; 5 with pulmonary atresia and 2 with absent pulmonary valve) were studied. Pulmonary RF was 28.3 ± 13.4%. Indexed RVEDV was 132 ± 33 mL/m^2^. Ejection fraction of RV and LV was 50.3 ± 7.8% and 59.1 ± 6.1%, respectively. Peak oxygen consumption (pVO2) was 71 ± 16% of predicted maximum value. A strong positive correlation was noted between CMR data including RVEDV, RVSV and LVSV, and pVO2. Higher RVEDV was correlated with higher RVSV and LVSV and higher pVO2, whereas lower RVEDV was associated with lower RVSV and LVSV and lower pVO2.

**Conclusion:** In rTOF, smaller RV resulted in reduced SV of both ventricles and significantly lower pVO2, whereas larger RV provided higher SV and higher pVO2 regardless of RVEF or RF. Smaller RV in rTOF may represent a unique pathological entity responsible for reduced exercise performance, which requires special consideration when determining further surgical interventions.

**Clinical Perspective:** 

**What is New?:** We characterized a novel clinical entity after surgical repair of tetralogy of Fallot (TOF) with a relatively small right ventricle (RV) and decreased exercise performance. It is likely due to limited RV stroke volume adjustment in response to peak exercise affecting left ventricular (LV) stroke volume. Although the pathogenesis of this smaller RV remains undetermined, our results shed light on the diverse clinical phenotypes after surgical repair of TOF.

**What are the Clinical Implications?:** Pulmonary valve replacement (PVR) is a treatment option for progressive RV dilatation and/or symptoms of exercise intolerance related to persistent pulmonary valve insufficiency in repaired TOF. Our data demonstrated that poor exercise performance was more frequently associated with a smaller RV size rather than dilated RV. Indication for PVR in repaired TOF needs to be carefully assessed in symptomatic patients with non-enlarged RV.

## Introduction

Persistent right ventricular (RV) volume overload can cause RV dilatation and dysfunction resulting in RV failure, decreased exercise tolerance and quality of life, and increased premature mortality. This condition is frequently encountered in several congenital heart diseases (CHD) including tetralogy of Fallot (TOF) following transannular patch repair, residual pulmonary valve insufficiency (PI) following balloon valvuloplasty for severe or critical congenital valvar pulmonary stenosis (PS), and after creation of right ventricular outflow tract (RVOT) by a conduit in CHD with certain conotruncal anomalies or after Ross operation [1].

Pulmonary valve replacement (PVR) is a treatment choice for progressive RV dilatation due to persistent PI, but the optimum timing of PVR remains controversial because of its limited longevity [2–4]. Currently, measurement of RV volume estimated by cardiac magnetic resonance (CMR) imaging is regarded as a gold standard to determine the timing of PVR as excessive RV dilatation may not be recoverable [5–7]. Symptoms including exercise intolerance or exercise-induced discomfort are also recognized as important indicators for PVR [8]. However, a correlation between the degree of RV dilatation and exercise-induced symptoms has been inconsistent.

As a noninvasive technique to gauge the level of fitness in patients with CHD, cardiopulmonary exercise testing (CPET) has the capability to assess peak cardiac output, stroke volume (SV), pulmonary function through oxygenation and ventilation, and peripheral muscular status [9]. An indication of CPET in CHD patients includes evaluation of exercise-induced symptoms and arrythmias in addition to quantification of cardiopulmonary reserve [10]. For patients with repaired TOF (rTOF) who subsequently may need PVR, preoperative CPET has been proposed as routine assessment of predictor of surgical outcome, where peak oxygen consumption (pVO2) was found to have the strongest predictor of early mortality with PVR on multivariable analysis [11]. However, the relationship between hemodynamic status and exercise performance has been poorly understood in rTOF.

A primary aim of this study was to investigate whether CPET represents the degree of RV dilatation and dysfunction secondary to chronic volume overload induced by persistent PI in rTOF patients. We aimed to explore how CMR volumetric data correlate with peak exercise performance measured by CPET.

## Methods

This retrospective study was approved by the Nemours Children’s Health Institutional Review Board in Wilmington, Delaware. The study protocol conformed to the ethical guidelines established by the 1975 Declaration of Helsinki.

### Patients

We enrolled patients with rTOF who underwent surgical repair during infancy and who had both CMR and CPET after 10 years of age. Patients with genetic syndromes involving multiple organ systems, those with intellectual disability, or obese individuals (body mass index > 30 kg/m^2^) were excluded. Patients with obesity were excluded as it complicates the interpretation of CPET [12]. Only those who reached peak exercise level were included, either peak respiratory exchange ratio (RER) ≥ 1.01 or > 90% of estimated maximum heart rate (HR) for age (= 220 - Age). Demographic information, diagnosis of primary heart disease, and surgical and cardiac catheterization history were obtained from electronic medical health records. The patients’ medical information was tightly protected according to Health Insurance Portability and Accountability Act regulation to maintain patient confidentiality and safety with de-identified data.

### CPET

All study subjects underwent symptom-limited bike CPET consisting of an electronically braked cycle ergometer following the RAMP (Raise, Activate, Mobilize, and Potentiate) protocol with work rate (WR) of approximately 0.3 Watts/minute increment. Heart rate and oxygen saturation (SaO2) were monitored continuously, and blood pressure (BP) was recorded every 2-3 minutes throughout the study and at peak exercise. A 12-lead electrocardiogram (ECG) was recorded continuously throughout the study. Work rate, minute ventilation (VE), oxygen consumption (VO2), and carbon dioxide production (VCO2) were measured continuously. Peak and submaximal CPET parameters were obtained. Peak parameters included pVO2, peak oxygen pulse (pOP), peak work rate (pWR), peak VE, and peak respiratory exchange ratio. Peak VO2 was also presented as %predicted maximum VO2 (%PmaxVO2). Submaximal parameters were obtained including ventilatory anaerobic threshold (VAT), a slope of VO2 and HR (ΔVO2/ΔHR; surrogate of SV), and a slope of VE and VCO2 (ΔVE/ΔVCO2; ventilatory efficiency or ventilatory equivalent of carbon dioxide). Ventilatory anaerobic threshold was also presented as %VAT/PmaxVO2. Both absolute and weight-indexed values were presented when appropriate.

### CMR

Cardiac magnetic resonance imaging examinations were performed on a 1.5-Tesla MRI scanner (GE Medical System, Chicago, IL United States or Siemens, Germany). Sequences to assess anatomy, volume, function, and phase contrast imaging were reviewed. Obtained parameters include RV and left ventricular (LV) ejection fractions (EF) (RVEF and LVEF) (%), end-diastolic volume (RVEDV and LVEDV) (ml), SV (RVSV and LVSV) (ml), and pulmonary regurgitant fraction (PRF) (%). Ventricular volumes were also expressed as body surface area (BSA)-indexed values (RVEDVi, LVEDVi, RVSVi, and LVSVi) (ml/m^2^) with corresponding z-scores [13] .

### Statistics

Descriptive statistics were represented as mean with standard deviation and/or range.

Linear regression curves with Spearman correlation were used to evaluate independent correlations. Kruskal Wallis test (nonparametric analysis of variance) with Dunn’s multiple comparison. Statistical significance with Student t-test was defined as *p*-value < 0.05. The statistical software GraphPad Prism 6 was used for data analysis (San Diego, CA; https://www.graphpad.com).

## Results

### Subtypes of TOF and Surgical Procedures

Patient demographics and clinical characteristics are summarized in **Table 1**. The average age of patients enrolled in CPET was 18.3 ± 4.7 years, and they were predominantly females (62%). The primary diagnosis was TOF with pulmonary stenosis (81%), followed by TOF with pulmonary atresia (14%), and TOF with absent pulmonary valve (5%). Initial reparative surgical intervention was predominantly transannular patch repair (89%); three patients initially received an RV to pulmonary artery (PA) homograft, and one patient underwent creation of a monocusp valve. Six patients required a palliative intervention prior to primary surgical repair of TOF. The average age of initial surgical repair was 1.8 months. After this combined CMR and CPET assessment, two patients underwent RVOT revision, and five patients had PVR.

**Table 1.**
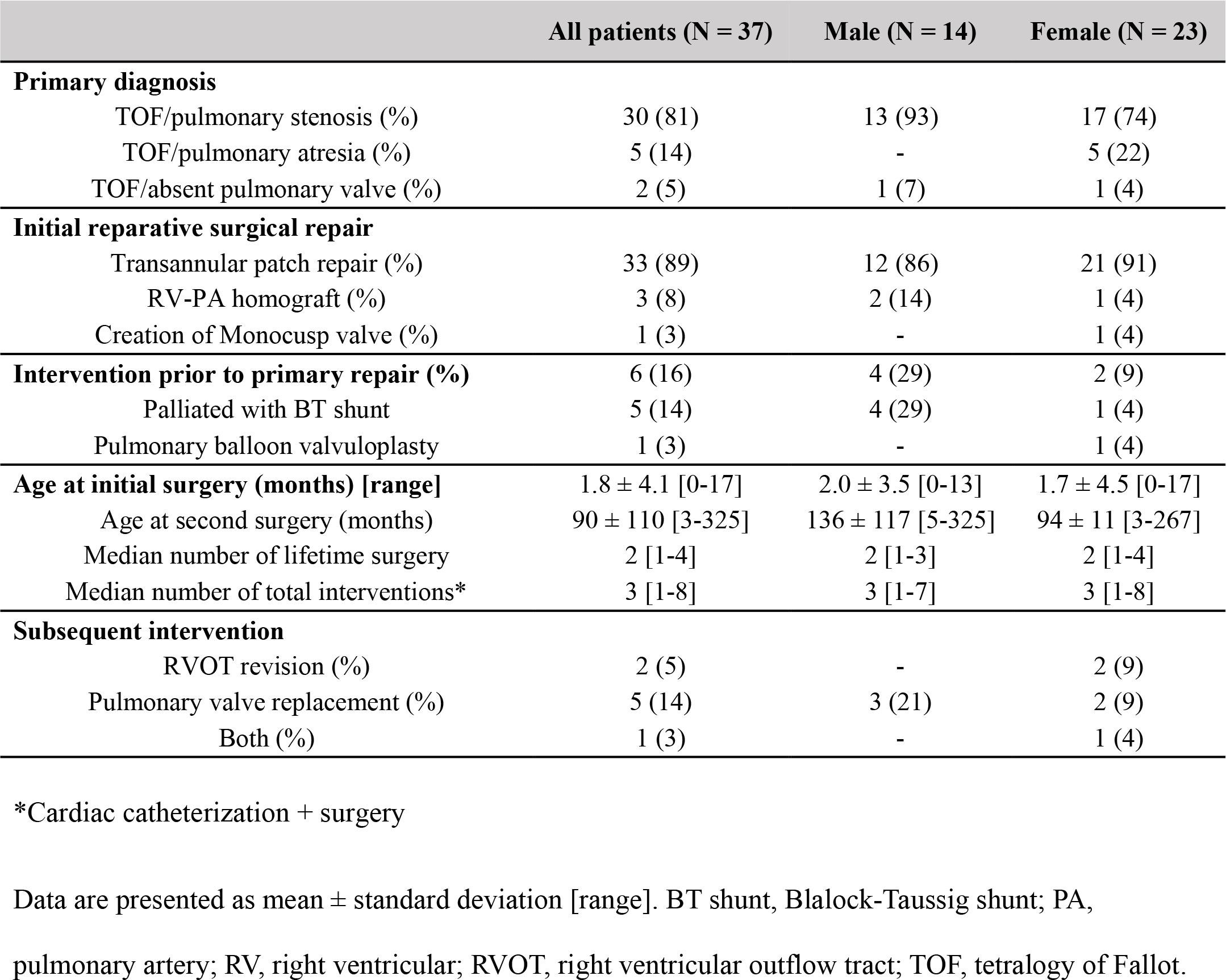
Patients’ Background Information.

### CPET Data

The CPET data including peak and submaximal values are summarized in **Table 2**. Males and females are presented separately as there are significant differences in CPET values by sex [14]. Weight and height were significantly higher in males, but BMI was comparable. Males showed significantly higher peak systolic BP, pVO2/kg, pOP, pWR, pWR/kg, and pVE than females. However, %PmaxVO2 was comparable between males and females (approximately 70%), which was mildly decreased when compared with the reference values, consistent with the published studies [15–17]. The %VAT/PmaxVO2 was also comparable between males and females. Males showed higher ΔVO2/ΔHR (both absolute and weight-indexed values) and lower ΔVE/ΔVCO2, suggesting higher SV and better ventilatory efficiency than females. These sex differences are comparable with those in the normal population except for ΔVE/ΔVCO2 [14].

**Table 2.** CPET Parameters.

CPET Parameters
|  | All patients (N = 37) | Male (N = 14) | Female (N = 23) | *p Value* |
| --- | --- | --- | --- | --- |
| **Patient demographics** |  |  |  |  |
| Age at CPET (years) | 18.3 ± 4.7 | 20.0 ± 6.0 [13-30] | 17.3 ± 3.5 [12-25] |  |
| Weight (kg) | 66.3 ± 12.7 [39.7-99.5] | 74.4 ± 12.8 [53.7-99.5] | 61.4 ± 9.9 [39.7-78] | **0.0015** |
| Height (cm) | 167 ± 11 [144-187] | 175 ± 4 [169-185] | 162 ± 10 [144-187] | **< 0.0001** |
| BMI (kg/m<sup>2</sup>) | 23.9 ± 3.8 [16-32] | 24.2 ± 4.2 [17-32] | 23.6 ± 3.6 [16-30] | 0.679 |
| **Peak CPET values** |  |  |  |  |
| Peak HR (bpm) | 175 ± 19 [130-205] | 178 ± 23 [134-205] | 173 ± 17 [130-196] | 0.392 |
| Peak SBP (mmHg) | 150 ± 23 [108-200] | 162 ± 22 [108-196] | 142 ± 20 [118-200] | **0.007** |
| pVO<sub>2</sub>(mL/kg/min) | 27.5 ± 6.8 [17.6-42.4] | 30.7 ± 7.7 [19.1-42.4] | 25.5 ± 5.5 [17.6-38] | **0.023** |
| pVO<sub>2</sub>/predicted max VO<sub>2</sub> (%) | 71 ± 16 [42-109] | 68.1 ± 17.1 [42.4-94.2] | 73 ± 15.6 [50-109] | 0.389 |
| pOP (mL/beat) | 10.5 ± 2.8 [5.4-17.1] | 12.9 ± 2.4 [8.9-17.1] | 9.0 ± 1.8 [5.4-14.1] | **< 0.0001** |
| pWR (Watts) | 131 ± 40 [51-208] | 163 ± 34 [113-208] | 111 ± 29 [51-180] | **< 0.0001** |
| pWR (Watts/kg) | 1.97 ± 0.51 [1.23-3.12] | 2.25 ± 0.57 [1.41-3.12] | 1.81 ± 0.40 [1.23-2.65] | **0.009** |
| pVE (L/min) | 77.7 ± 24.5 [33.2-154.7] | 93.8 ± 27.8 [63.7-154.7] | 67.9 ± 16.6 [33.2-106.4] | **0.001** |
| pRER | 1.20 ± 0.11 [1.01-1.62] | 1.26 ± 0.12 [1.13-1.62] | 1.20 ± 0.10 [1.01-1.42] | 0.144 |
| **Submaximal CPET values** |  |  |  |  |
| VAT (mL/kg/min) | 20 ± 7.5 [9.5-48] | 22.5 ± 10.4 [10.6-48] | 18.6 ± 4.9 [9.5-32] | 0.142 |
| VAT/predicted max VO<sub>2</sub> (%) | 51 ± 19 [24-107] | 50 ± 23 [24-107] | 53 ± 14 [27-92] | 0.595 |
| ΔVE/ΔVCO<sub>2</sub> | 28.2 ± 6.1 [18.8-50.3] | 25.4 ± 3.8 [18.8-31.3] | 30.0 ± 6.6 [21.2-50.3] | **0.023** |
| ΔVO<sub>2</sub>/ΔHR | 16.3 ± 6.41 [6-31.1] | 20.1 ± 6.8 [8-31.1] | 13.9 ± 4.95 [6-27.3] | **0.003** |
| Δ[VO<sub>2</sub>/kg]/ΔHR | 0.24 ± 0.09 [0.09-0.49] | 0.28 ± 0.11 [0.09-0.49] | 0.21 ± 0.06 [0.09-0.34] | **0.018** |

Data are presented as mean ± standard deviation [range]. P < 0.05 indicates statistical significance.
BMI, body mass index; CPET, cardiopulmonary exercise testing; HR, heart rate; pOP, peak oxygen pulse; pRER, peak respiratory exchange ratio; pVE, peak minute ventilation; pVO<sub>2</sub>, peak oxygen consumption; pWR, peak work rate; SBP, systolic blood pressure; VAT, ventilatory anaerobic threshold; VE, minute ventilation; VCO<sub>2</sub>, carbon dioxide production; VO<sub>2</sub>, oxygen consumption.

### Volume Measurement by CMR

Volumetric data by CMR are summarized in **Table 3**. Both RVEDVi and RVSVi were significantly increased in males and females, whereas LVEDVi and LVSVi were not increased.

**Table 3.** CMR Data.

| CMR parameters | All patients (N = 37) | Male (N = 14) | Female (N = 23) | *p* Value |
| --- | --- | --- | --- | --- |
| PRF (%) | 28.1 ± 13.9 | 26.1 ± 15.7 | 29.0 ± 13.1 | 0.498 |
| RVEF (%) | 49.8 ± 8.0 [23-66] | 46.9 ± 9.3 [23-65] | 51.0 ± 6.6 [40-66] | 0.094 |
| LVEF (%) | 58.4 ± 7.1 [39-75] | 57.3 ± 7.5 [42-75] | 59.0 ± 6.9 [39-75] | 0.484 |
| RVEDV (mL) | 226 ± 58 [92-352] | 267 ± 46 [186-353] | 201 ± 50 [92-315] | **0.0003** |
| RVEDVi (mL/m<sup>2</sup>) | 127 ± 28 [72-189] | 142 ± 24 [112-189] | 118 ± 27 [72-185] | **0.009** |
| **RVEDV z score** | **2.55 ± 1.77 [-1.77-6.08]** | **2.39 ± 1.34 [0.43-5.33]** | **2.64 ± 1.96 [-1.77-6.08]** | 0.696 |
| LVEDV (mL) | 141 ± 38 [69-234] | 161 ± 37 [98-220] | 130 ± 34 [69-234] | **0.013** |
| LVEDVi (mL/m<sup>2</sup>) | 82 ± 17 [55-137] | 87 ± 16 [63-117] | 78 ± 17 [55-137] | 0.118 |
| LVEDV z score | -0.67 ± 1.85 [-3.66-3.75] | -0.97 ± 1.60 [-3.66-2.11] | -0.51 ± 1.96 [-3.40-3.75] | 0.488 |
| RVSv (mL) | 111 ± 32 [52-189] | 125 ± 30 [67-170] | 103 ± 31 [52-189] | **0.044** |
| RVSVi (mL/m<sup>2</sup>) | 65 ± 16 [32-111] | 68 ± 15 [34-90] | 63 ± 16 [32-111] | 0.291 |
| RVSv z score | 1.10 ± 1.89 [-5.15-4.63] | 0.36 ± 2.08 [-5.15-3.08] | 1.50 ± 1.65 [-2.56-4.63] | 0.087 |
| LVSv (mL) | 81 ± 20 [50-126] | 92 ± 23 [59-126] | 75 ± 15 [50-104] | **0.009** |
| LVSVi (mL/m<sup>2</sup>) | 47 ± 9 [32.0-70.5] | 50 ± 11 [32.0-71] | 45 ± 7 [35-59] | 0.115 |
| LVSv z score | -1.24 ± 1.52 [-5.81-2.11] | -1.73 ± 1.78 [-5.81-1.49] | -0.97 ± 1.29 [-3.12-2.11] | 0.151 |

Data are presented as mean ± standard deviation [range]. *p* < 0.05 indicates statistical significance.
CMR, cardiac magnetic resonance; LVEDV, left ventricular end-diastole volume; LVEDVi, indexed left ventricular end-diastole volume; LVEF, left ventricular ejection fraction; LVSv, left ventricular stroke volume; LVSVi, indexed left ventricular stroke volume; PRF, pulmonary regurgitant fraction; RVEDV, right ventricular end-diastole volume; RVEDVi, indexed right ventricular end-diastole

The pulmonary regurgitant fraction was comparable between males and females. Absolute volume values were significantly higher in males than in females, including RVEDV, LVEDV, RVSV, and LVSV, most likely reflecting the difference in body size. The BSA-indexed values, LVEDVi, RVSVi, and LVSVi, were comparable between males and females, whereas RVEDVi was significantly higher in males than in females. There were no statistically significant differences between male and female volumetric z scores. On average, the RVEDV z scores were higher in rTOF patients (2.55 ± 1.77). Z scores of LVEDV, RVSV, and LVSV were comparable with normal reference ranges.

### Progression of RV Remodeling and Exercise Performance

To understand the progression of RV dilatation and dysfunction in response to persistent RV volume overload, we arbitrarily divided the cohort into four groups by the degree of RV dilatation (threshold of RVEDVi at 125 mL/m^2^) and RV systolic function (threshold of RVEF at 55%) (**Figure 1*a***). Group A represents no significant RV dilatation with preserved RV systolic function (RVEDVi < 125 mL/m^2^ and RVEF > 55%). Group B represents those with significant RV dilatation with preserved RV systolic function (RVEDVi ≥ 125 mL/m^2^ and RVEF > 55%). Group B is regarded as a slightly advanced stage of RV volume overload from Group A. Group C represents further progression from B with RV dilatation and RV systolic dysfunction (RVEDVi ≥ 125 mL/m^2^ and RVEF < 55%). Group D refers to a group with decreased RV systolic function without RV dilatation, defined as RVEDVi < 125 mL/m^2^ and RVEF < 55%.

**Figure 1.**
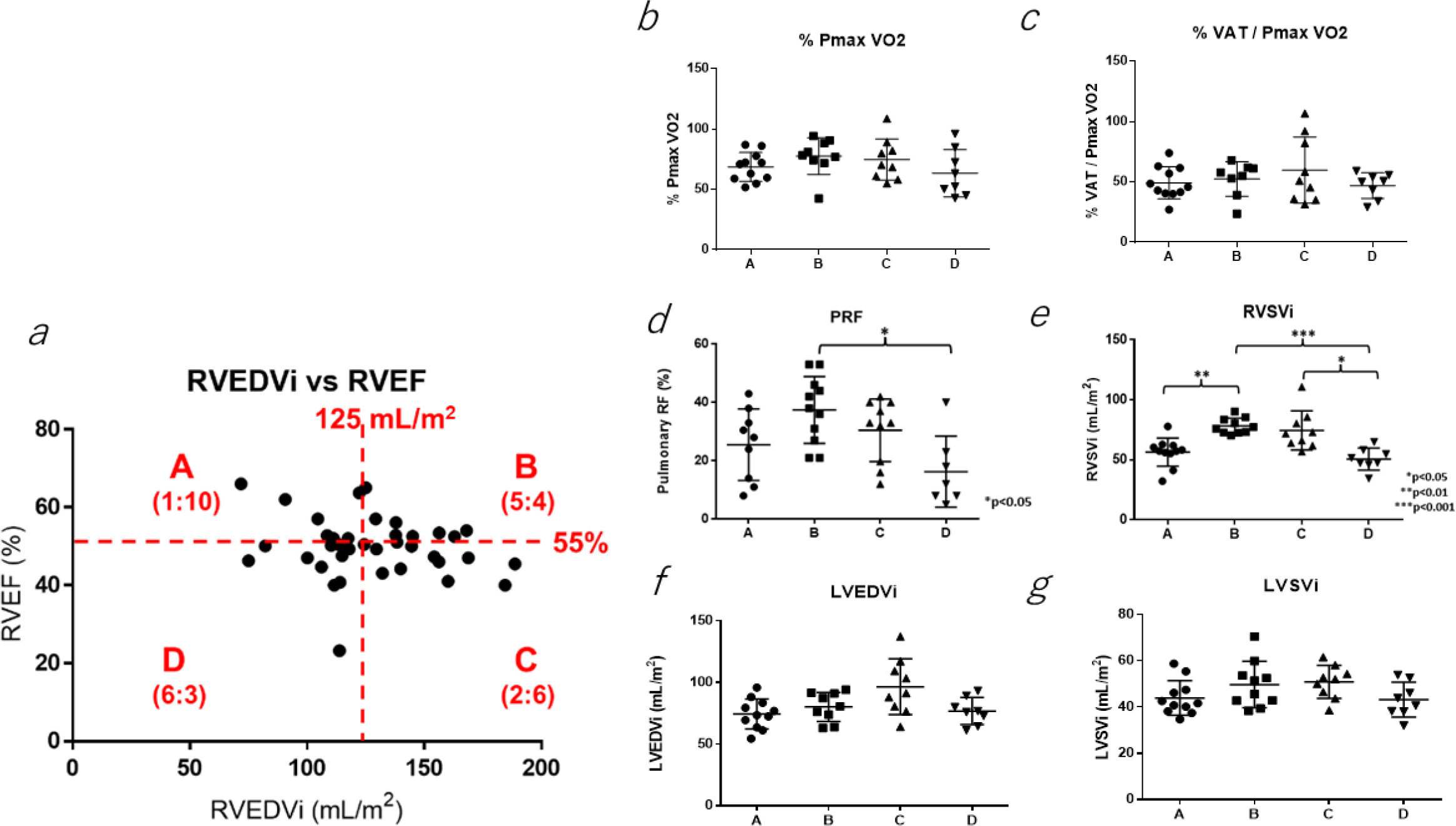
***a***: Sequence of right ventricular (RV) remodeling in response to chronic pulmonary insufficiency. Patients were divided into four groups by indexed right ventricular end-diastole volume (RVEDVi) of 125 ml/m^2^ and right ventricular ejection fraction (RVEF) of 55%. The levels of RV remodeling were graded as A (less-dilated RV with normal systolic function: mild), B (dilated RV with normal systolic function: moderate), and C (dilated RV with decreased systolic function: advanced). D is a special group with less dilated RV with decreased systolic function. Sex distribution is shown as (male : female). Certain CPET and CMR parameters were compared within the four groups. **b**: %PmaxVO2 (%), **c**: %VAT/PmaxVO2 (%), **d**: PRF (%), **e**: RVSVi (ml/m^2^), **f**: LVEDVi (ml/m^2^), and **g**: LVSVi (ml/m^2^). LVEDVi, indexed left ventricular end-diastole volume; LVSVi, indexed left ventricular stroke volume; pVO2, peak oxygen consumption; %PmaxVO2, % predicted max VO2; PRF, pulmonary regurgitant fraction; RVSV1, right ventricular stroke volume; VAT, ventilatory anaerobic threshold.

Figures 1b to **1*g*** show certain CPET and CMR parameters in four groups. There was no significant difference between the four groups in %PmaxVO2, %VAT/PmaxVO2, LVEDVi, or LVSVi. More importantly, there was no decline in %PmaxVO2 or %VAT/PmaxVO2 in response to the degree of progress in RV remodeling (from groups A, B, to C). Group D exhibited significantly lower PRF than group B (*p* < 0.05). Larger RVEDVi groups (groups B and C) demonstrated significantly higher RVSVi than smaller RVEDV groups (groups A and D). Our data indicate that larger RVEDVi was associated with higher RVSVi regardless of RV systolic function. Both LVEDVi and LVSVi showed a similar trend as RVSVi in four groups but did not show statistical significance. In contrast, smaller RV groups (A and D) revealed significantly lower RVSVi than larger RV groups (B and C) regardless of systolic function.

### RV Dilatation is Associated with Higher SV and Better Exercise Performance

Correlations between CMR measurements and CPET values are shown in Figure 2. Non-indexed absolute values were compared. There was a statistically significant correlation between RVEDV and pVO2 (*p* = 0.0003), RVEDV and RVSV (*p* < 0.0001), RVSV and LVSV (*p* < 0.0001), RVSV and pVO2 (*p* < 0.0001), RVEDV and LVEDV (*p* < 0.0001), and LVSV and pVO2 (*p* = 0.0005). Our data indicate that larger RVEDV was associated with higher RVSV, LVSV, and pVO2, suggesting the larger RV size are associated with higher peak exercise performance; the degree of RV dilatation had nothing to do with low SV or low pVO2.

**Figure 2.**
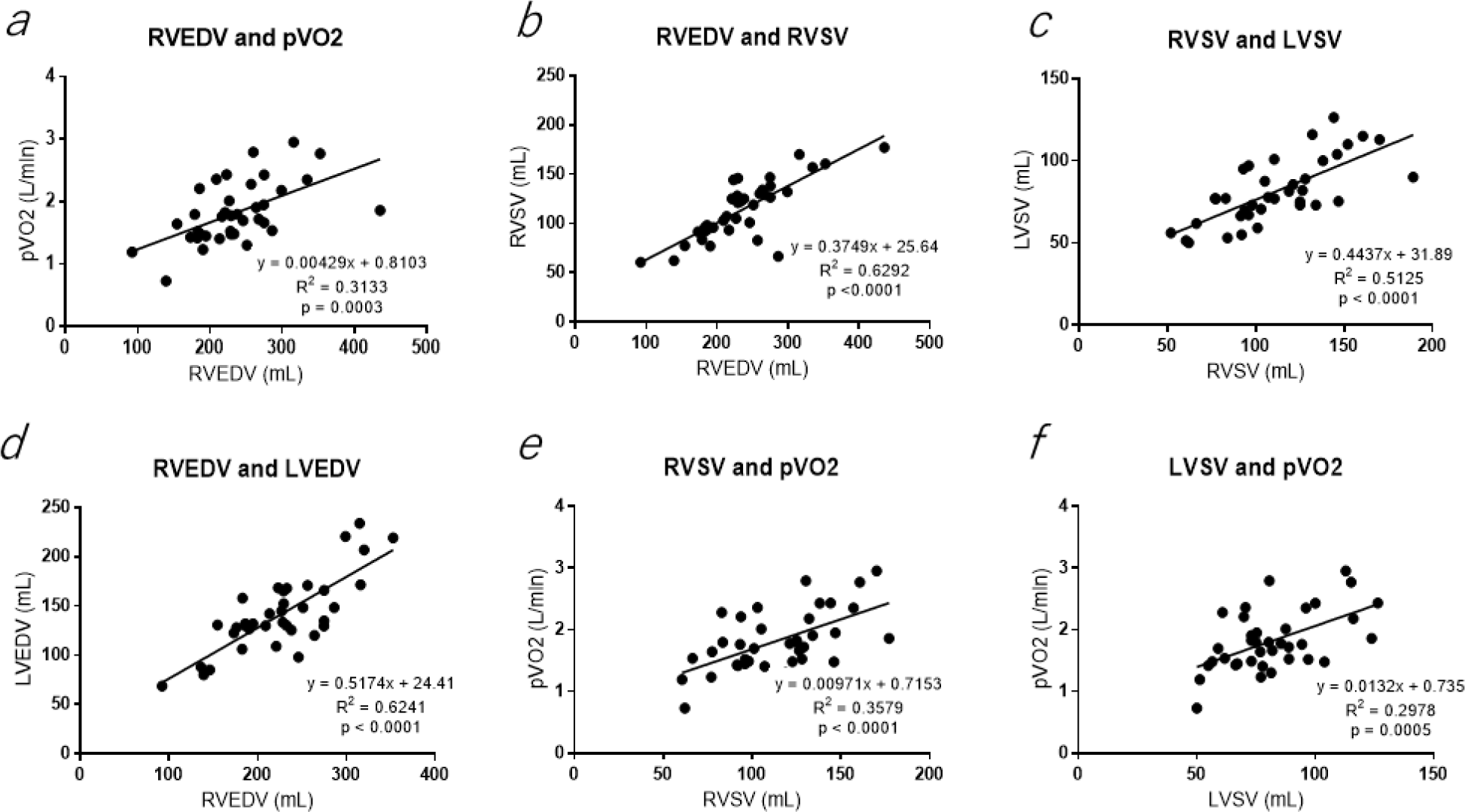
Correlations between parameters of cardiopulmonary exercise testing and cardiac magnetic resonance. ***a***: RVEDV (ml) and pVO2 (L/min), ***b***: RVEDV (ml) and RVSV (ml), ***c***: RVSV (ml) and LVSV (ml), ***d***: RVEDV (ml) and LVEDV (ml), ***e***: RVSV (ml) and pVO2 (L/min), and ***f***: LVSV (ml) and pVO2 (L/min). All showed an excellent positive correlation (p ≤ 0.0005). LVEDV, left ventricular end-diastole volume; LVSV, left ventricular stroke volume; pVO2, peak oxygen consumption; RVEDV, right ventricular end-diastole volume; RVSV, right ventricular stroke volume.

Because there were significant sex differences in CPET and CMR parameters (**Tables 2** and **3**), the correlation curves obtained in Figure 2 were expressed males and females separately (Figure 3), which showed the same trends seen in Figure 2 except no strong correlation between RVEDV and pVO2 in males (Figure 3a). It was a smaller RV that revealed significantly lower SV and lower pVO2 in both males and females.

**Figure 3.**
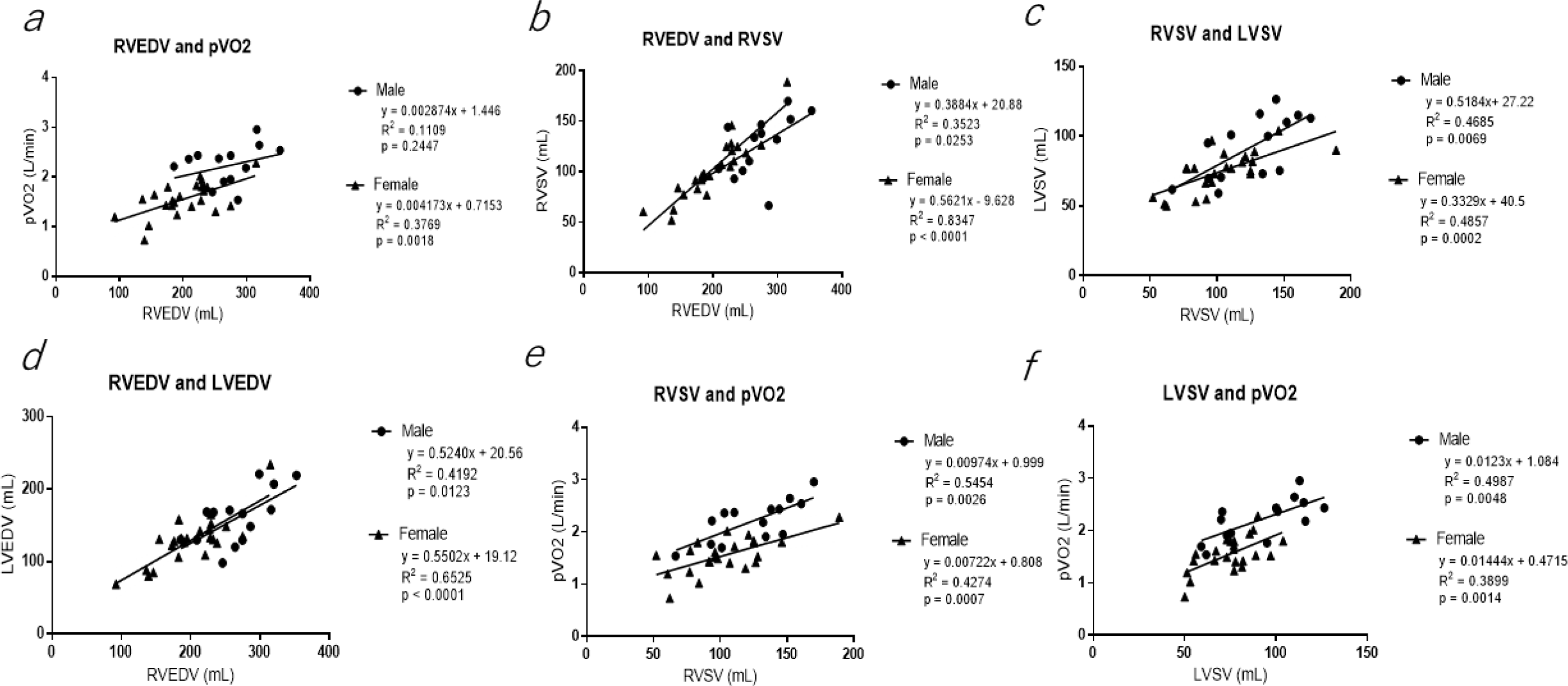
The correlation curves in Figure 2 are presented in male and female separately. a: RVEDV (ml) and pVO2 (L/min), b: RVEDV (ml) and RVSV (ml), c: RVSV (ml) and LVSC (ml), d: RVEDV (ml) and LVEDV (ml), e: RVSV (ml) and pVO2 (L/min), and f: LVSV (ml) and pVO2 (L/min). All showed significant positive correlation (p < 0.05) except RVEDV and pVO2 correlation in male (*a*). LVEDV, left ventricular end-diastole volume; LVSV, left ventricular stroke volume; pVO2, peak oxygen consumption; RVEDV, right ventricular end- diastole volume; RVSV, right ventricular stroke volume.

### Clinical Outcomes after PVR

Of the 37-patient cohort, a total of nine patients underwent PVR. Comparative data (symptoms, pVO2, and RVEDVi) of patients before and after PVR are summarized in **Table 4**. Only two patients were noted to have symptoms prior to PVR, and both continued to have symptoms afterward. Many patients developed symptoms following PVR. Of five patients who had CPET before and after PVR, none showed improvement in pVO2/kg. After PVR, RVEDVi decreased in three patients but increased in two patients. Two patients needed repeat PVR.

**Table 4.** Symptoms, %predicted pVO2, and RVEDVi before and after PVR.

| Pt | Prior to PVR |  |  | After 1st PVR |  |  | After 2nd PVR |  |  |
| --- | --- | --- | --- | --- | --- | --- | --- | --- | --- |
|  | Symptoms | pVO2/kg<br>(%predicted) | RVEDVi<br>(mL/m<sup>2</sup>) | Symptoms | pVO2/kg<br>(%predicted) | RVEDVi<br>(mL/m<sup>2</sup>) | Symptoms | pVO2/kg<br>(%predicted) | RVEDVi<br>(mL/m<sup>2</sup>) |
| 1 | None |  | 160 | None →<br>endocarditis | - | - | None | 29.4 (85) | 100 |
| 2 | Dizziness,<br>lightheaded,<br>shortness of<br>breath | 18.1 (52) | 110 | Chronic fatigue | - | - |  |  |  |
| 3 | None | 27.2 (53) | 190 | Exertional<br>dyspnea and<br>syncope | 19.2 (37) | 148 | None | 21.3 (49) | 117 |
| 4 | Dizziness | 20 (47) | 122 | Palpitations | - | - |  |  |  |
| 5 | None | 33.9 (69) | 94 | None | 30.8 (63) | 132 |  |  |  |
| 6 | None | 27.4 (65) | 138 | Vasovagal<br>syncope and<br>palpitations | - | - |  |  |  |
| 7 | None | 48.9 (92) | 188 | None | 36.4 (86) | 169 |  |  |  |
| 8 | None | 47 (90) | 165 | Chest pain | 38 (80) | 185 |  |  |  |
| 9 | None | 40.1 (87) | 189 | Palpitations | 34.3 (76) | - |  |  |  |

pVO2, peak oxygen consumption; PVR, pulmonary valve replacement; RVEDVi, right ventricular end-diastolic volume indexed by body surface area.
Unit of pVO2/kg is ml/kg/min.

## Discussion

Our current data showed that patients with rTOF demonstrated mildly diminished peak exercise performance (approximately 70% of predicted max VO2) in both males and females. Surprisingly, the severity of RV dilatation indicated by RVEDV (ml) had no correlation with decreased exercise performance. On the contrary, there was a significant positive correlation between RVEDV and peak exercise performance presented by peak VO2 (L/min) in both males and females. An enlarged RV provided higher RVSV and LVSV and thus higher pVO2 than smaller RV. Patients with smaller RV revealed poorer exercise performance regardless of systolic function. Our findings suggest complex changes in RV myocardium after surgical repair of TOF, responsible for diverse clinical phenotype in rTOF.

### Development of RV Remodeling and Exercise Performance

Our data demonstrated that pVO2 did not decrease by the degree of RV dilatation. In fact, larger RV provided higher SV (both RV and LV) and better pVO2 than smaller RV (Figures 2 and **3**). Roest et al. performed a prospective CMR study of 15 patients with rTOF at rest and submaximal exercise (∼ 60% peak level) and demonstrated that the degree of PI decreased significantly during supine bike exercise (*p* = 0.012), suggesting PI and RV dilatation may not be a strong determinant of exercise intolerance [18]. Our study revealed that the smaller RV was correlated with lower RVSV, lower LVSV, and lower pVO2 regardless of RV systolic function (groups A and D in comparison with groups B and C in Figure 2); both RVSV and LVSV at rest by CMR correlated well with pVO2, as also shown in a recent study [16].

O’Meagher et al. compared rTOF patients between higher RVEDVi (≥ 150 ml/m^2^) and lower RVEDVi (< 150 ml/m^2^) and showed that there were no differences in LVEF, LVSVi, or exercise capacity including VAT/kg and pVO2/kg [19]. In another study of rTOF patients who underwent CPET and CMR, RVEF was the only CMR predictor of %Pmax VO2, pOP, and VAT [20]. It was proposed from a recent study that RVEF < 40% may be useful to identify prognostically significant reduction in exercise capacity with varying degrees of RV dilatation [21]. In our study, there was only one patient with RVEF < 40% (see Figure 1A). Our data as well as other published studies indicate that peak exercise performance and exercise-induced symptoms do not represent the degree of RV dilatation or dysfunction [22].

### Small RV Despite PI, a New Pathological Entity in rTOF?

Our study demonstrated that patients with lower RVEDVi (smaller RV) had significantly lower RVSVi regardless of systolic function (Figure 1e). There was strong positive correlation between RVSV and pVO2 (both absolute values; Figure 2e). It is plausible that small RV represents hemodynamically restrictive ventricle causing diastolic dysfunction, which impairs diastolic relaxation and filling and limits cardiac output increase during exercise [23]. In our cohort, there were no differences in surgical profile or number of surgeries between smaller and larger RV groups (**Suppl. Table 1**). The amount of PI and consequent RV dilatation and dysfunction are regarded as major prognostic determinants following TOF repair. It has been proposed that the presence of a restrictive RV limits the detrimental effects of PI that enhances better antegrade flow across RVOT and improve exercise performance [24], which is completely opposite from our findings of smaller RV having worse exercise performance. Regarding CPET analysis in rTOF, small RV has never been addressed as a cause of poor exercise performance in the recent review article [25]. The pathogenesis of small RV in rTOF, however, remains undetermined in this study.

Kim et al. studied 48 rTOF patients and demonstrated that those with higher pulmonary artery elastance (or advanced pulmonary vascular dysfunction) were associated with higher mean PA pressure, smaller RVEDVi, and significantly lower pVO2, describing a similar clinical entity of small RV size with poor exercise performance [26]. They suggested decreased pulmonary vascular distensibility as a cause of impaired exercise performance in rTOF, probably due to suppression of increased antegrade flow across an incompetent pulmonary valve during exercise. The authors did not assess the direct relationship between RV size and pVO2. The interaction between RV and PA (RV-PA coupling) may be another important determinant of effective SV and exercise performance in rTOF [27]. In older patients (33 ± 13 years), Egbe et al. demonstrated a similar finding of negative correlation with PA elastance and exercise capacity [28]. However, the relationship between smaller RV and increased PA elastance was not discussed in these studies.

### Indication and Optimum Timing for PVR

The optimum timing of PVR in rTOF patients has been extensively discussed [29–32]. The controversy of indications and optimum timing of PVR is primarily because of the limited longevity of the prosthetic valve. Valve function in pediatric patients with CHD, mainly TOF, who undergo valve replacement in the pulmonary position can be expected to remain stable for 5 years after placement [33], but this may be shorter in younger patients. By 10 years, most patients (approximately 80%) will develop valvular dysfunction and often require reoperation. In another study of pediatric rTOF patients who underwent PVR, freedom from reoperation was approximately 66% and 21% at 10 and 15 years, respectively [34]. The challenge becomes when determining the need for PVR to prevent irreversible RV remodeling against the timed life expectancy of prosthetic valves that lead to subsequent reoperations. The timing of PVR is usually determined by the degree of RV dilatation and/or the presence of exercise-related symptoms [35]. The RV volume measurement by CMR has become a gold standard to determine the indication for PVR for rTOF; patients with RVEDVi > 150-180 ml/m^2^ are indicated for PVR as RV dilatation may not recover once RVEDVi exceeds more than 170 ml/m^2^ [4,35,36].

Symptomatic patients with exercise intolerance not explained by extracardiac causes or chronotropic incompetence are often considered for PVR [35]. The CPET is routinely performed to assess the degree of exercise intolerance for the indication for PVR. However, the CPET parameters usually do not represent the severity of RV dilatation or dysfunction in rTOF as we presented in this study. In a recent meta-analysis of over 700 patients with rTOF, CPET values did not significantly change following PVR [37]. A few other studies also demonstrated that exercise performance did not improve after PVR for severe RV dilatation due to chronic PI despite significant reduction of RV dilatation [38–40], suggesting that peak exercise performance represented by pVO2 may be an independent parameter from the degree of RV dilatation.

### What Does PVR Do for rTOF with RV Dilatation?

Surgical indications for PVR for rTOF include progressive RV remodeling (RV dilatation and/or RV dysfunction), poor exercise performance, and symptoms with or without exercise. In a meta-analysis of 48 studies including over 3000 patients with rTOF who subsequently underwent PVR, there was noted improvement in biventricular function, RV volume, and symptoms [3]. Of these, 22 studies reported data on RVEDVi before and after PVR, and a statistically significant overall reduction in RVEDVi was noted. Twenty-six studies reported the New York Heart Association functional classification (NYHA) and demonstrated an overall significant reduction in NYHA classification following PVR. By the meta-regression method, patients with larger RV preoperatively had the best response in terms of RV geometry and decrease in RV volume postoperatively but had the lowest improvement in symptoms.

Although improvement of RV dilatation after PVR for advanced RV dilatation is quite consistent [41], the improvement of symptoms or exercise capacity by PVR remains controversial. Gengsakul et al. studied the outcome of PVR in adult rTOF patients and found that PVR resulted in subjective improvement in functional NYHA status and reduction in RVEDVi without significant improvement in exercise capacity [42]. Lack of improvement in objective CPET data was attributed to the presence of persistent RV dysfunction following PVR. Ho et al. studied CPET of rTOF (average age 20.0 years) before and after PVR and demonstrated no notable improvement in pVO2 despite significant reduction of RVEDVi by CMR [17]. It appears that the degree of RV remodeling, exercise performance, and symptoms do not always correlate well with one another.

### Limitations

We acknowledge several limitations in our retrospective, single-center study. First, there is an intrinsic selection bias since there were no standardized guidelines for timing of CPET and CMR in our cohort. Most of our patients underwent these diagnostic tests in consideration for PVR because of echocardiographic findings suggestive of RV remodeling and/or presence of exercise-related symptoms. Second, RV diastolic dysfunction was not assessed in this study as echocardiographic assessment of diastolic function was not consistently obtained. The assessment of RV diastolic function could provide an additional insight into the pathophysiology of smaller RV patients exhibiting worse exercise performance. Third, RV remodeling consisting of dilatation and dysfunction is a complex pathological process in rTOF. Our cohort consists of relatively young rTOF subjects, which may be quite different from adults with chronic RV dilatation. Advanced RV remodeling seen adult rTOF patients with myocardial fibrosis in addition to severe RV dilatation and dysfunction may present with additional pathological features. Additionally, CPET is affected by physical conditioning and body habitus, which could affect pVO2. These factors were not controlled in our study. Lastly, this is a retrospective, single-center study with a relatively small cohort size that limits statistical power.

## Conclusion

One of the chronic sequalae of surgically rTOF is progressive RV dilatation and dysfunction due to persistent PI, which is a common indication for PVR. Presence of exercise-induced symptoms and/or exercise intolerance is regarded as another relevant indication for surgical intervention. In our small cohort of rTOF patients, there was no correlation between the degree of RV dilatation and worsening of pVO2; on the contrary, the patients with more dilated RV tended to show better pVO2 than those with smaller RV regardless of systolic function. A group of rTOF patients presented with smaller RV volume with poor exercise performance, which may be a unique pathological entity that requires special consideration for further surgical management. Although our study highlights the diverse clinical phenotype of rTOF, the underlying pathogenesis of rTOF with small RV remains undetermined. Further clinical investigation is warranted to delineate pathophysiology of post-TOF patients, particularly in view of RV diastolic dysfunction and impaired RV-PA coupling.

## Data Availability

Data will be available upon contact with the corresponding author.

## Acknowledgements

Authors would like to acknowledge Gina D’Aloisio, MS for querying the exercise database and Kimberley Eissmann for editing the manuscript text.

## Data Availability Statement

Data will be available upon contact with the corresponding author.

